# Chronotype distinctness a key risk factor for emotional and cognitive disruption in shift working nurses

**DOI:** 10.64898/2026.08.21.26361010

**Authors:** Ashleigh Davies, Robert Hickman, Ziyuan Cai, Daniel Jie Lai, Adam Hampshire, Peter J. Hellyer, Sukhi Shergill, Teresa D’Oliveira

## Abstract

The prevalence of shift work-centric industries and the rise of flexible working render it crucial to investigate the health risks posed by unnatural patterns of work and sleep. Shift working in healthcare has been linked to emotional dysregulation, reduced alertness, and cognitive impacts, with Shift Work Disorder (SWD) classified as a circadian rhythm sleep-wake disorder in the DSM-5. These impacts have historically been attributed to schedule-related sleep disturbances; however certain individuals appear more acutely affected. One measure of individual inflexibility to altered routines is chronotype distinctness, an amplitude dimension of the Caen Chronotype Questionnaire (CCQ). Here, we use Structural Equation Modelling (SEM) to unravel the mechanisms by which sleep and chronotype distinctness govern the neuropsychiatric symptom profiles of a cohort of National Health Service (NHS) shift workers (n=102). We constructed a measurement model, extracting latent constructs from questionnaires for sleep disturbances (PSQI), chronotype distinctness (CCQ), mood disorder (MDQ), depression (PHQ8) and emotional reactivity (ERS), and cognitive assessments. Correlations were identified between constructs, then translated into two SEMs - day and rotating shifts respectively - with sleep and distinctness as predictors of detrimental effects. Models were tested for direct effects, significant paths, and overall model fit. We found that, whilst sleep governed fatigue-based symptoms in day-shift workers, chronotype distinctness determined the severity of adverse effects in rotating-shift workers, including mood disorders and cognitive impairments. We surmise that high chronotype distinctness should be considered a risk factor for adverse effects surrounding night and rotating-shift work, and that interventions should incorporate chronotype-specific remediation.

SUMMARY QUESTIONS

**What is already known on this topic:** It is understood that sleep and schedule disturbances caused by shift working have widespread detrimental effects on health including emotional dysregulation, reduced attention, alertness and cognitive function.

**What this study adds:** Here, we explore the role that a new measure of individual inflexibility – chronotype distinctness – may play in governing the detrimental effects of shift work, that have previously been attributed to sleep disturbances alone.

**How this study might affect research, practice or policy:** These findings suggest that chronotype distinctness should be considered as a personal risk factor for worse emotional and cognitive effects in shift workers; hence, workplace accommodations and interventions that consider chronotype may produce more effective remediative results.

## INTRODUCTION

Shift working is a key feature of the modern occupational landscape, associated with inherently shift-based vocations such as healthcare, emergency services and the transportation industry, but also characteristic of a modern approach to remote, flexible, and international working. Shift-based work involves around 20% of the global workforce [1], with close to 40% of UK workers employed by the NHS alone [2]. One large cohort study found that 12.7% of individuals involved in rotating shift work were diagnosed with Shift Work Disorder (SWD), recognised in the DSM-V as ‘circadian rhythm sleep-wake disorder - shift work type’, a condition encompassing the psychiatric symptoms associated with shift work: insomnia, excessive daytime sleepiness, persistent sleep disturbances, distress or impairment in social, occupational or other areas of functioning; those that had SWD were up to twice as likely to suffer from mood and affect disorders [3].

Circadian rhythms are biological rhythms governed by the Suprachiasmatic Nuclei (SCN), a cluster of brain neurons whose timing is governed by clock genes that are repeatedly expressed and self-inhibited [4]. This timing is used as a pacemaker for a whole set of biological processes, such as metabolism, hormone release, and the sleep-wake cycle [5]. These internal rhythms are entrained to daily cycles of light in order to optimise day-time activity, and, equally importantly, to enable night-time recuperation, consolidation and recovery [6]. However, shift working demands a biological timing that directly contradicts the natural timing of these circadian rhythms, expecting metabolism, hormone release and alertness at a time during which the body is biologically geared for rest, and vice versa. The resulting impact could be described as a form of occupational jet-lag.

One noticeable effect of this jet-lag is disruption to the timing, quality and biological efficacy of sleep, which can in turn lead to next day fatigue, reduced alertness, and impaired executive attention, increasing the likelihood of occupational hazards. Recent research into cognitive ‘readiness’ has identified chronic sleep deprivation as a form of cognitive insult, equivalent to that of a mild brain injury [7]. In the context of healthcare, where precision and efficiency are paramount, this reduced cognitive capacity poses a significant risk of harm to patients and staff alike [8]. As well as affecting alertness and job performance, poor sleep has been found to worsen next day mood and affect [9], with chronic exposure to disrupted sleep exacerbating this effect, and ultimately leading to increased risk of insomnia and mood disorders [10]. This effect is reflected in shift working populations by widespread increases in rates of depression, depressive symptoms, and anxiety [11]. Similar bidirectional relationships are found across sleep, cognitive function, mood, and affect; these mutually exacerbatory symptoms render the potential impacts of shift work concerning [12].

Research into the long-term detrimental effects of shift working is a relatively new field, with the majority of studies investigating single predictors of adverse outcomes, often focused around only shift work status or sleep disturbances,[13] hence, failing to capture the complex relationships between the latent constructs underpinning shift-work related health issues such as the exacerbatory effects of sleep, cognition and emotion on one another. We propose that the risk potential of this complex interplay of sleep, cognition and emotion as a combined individual profile has yet to be considered in research. Furthermore, research to date has largely focused on population-wide trends; even those that investigate temporal fluctuations in sleep-disturbance related symptoms treat them as a state-based issue rather than acknowledging the role of trait-based individual sensitivity [14]. Understanding the interactions between different predictors, and investigating individual sensitivity to shift working are key to reducing the harm caused.

Circadian research has recently identified an individual flexibility trait underpinning the extent to which an individual can adapt to varying schedules [15]. Understanding how these individual differences determine the nature and severity of symptom profiles highlights the relevance of a personalised risk assessment regarding adverse health outcomes. Chronotype refers to individual differences in circadian rhythm entrainment, caused by slight advances or delays in the timing of the SCN clock, leading to a tendency towards ‘morningness’ or ‘eveningness’; this can be assessed using the Caen Chronotype Questionnaire (CCQ). Chronotype has been linked to psychiatric health outcomes, with a higher prevalence of mood disorders associated with evening types in the shift working population [16], suggesting that the effect on mood arises from a mismatch between the biological clock and typical daily routine. An amplitude dimension has been derived from the CCQ - chronotype distinctness, which captures the extent of this individual inflexibility to altered or atypical routines [17]. High degrees of distinctness (i.e., low flexibility) have been linked to negative health outcomes, including reduced sleep quality, conscientiousness and general life satisfaction. Circadian flexibility has been found to depend on individual factors, for instance sex and general chronotype [18]. Shift workers have been found to have less rigid amplitude compared to day workers, which may be advantageous in tolerating shift work-related circadian misalignment [19].

Here, we explore how this chronotype distinctness measure could determine the severity of adverse health outcomes in the context of shift work. Considering distinctness as a trait-based measure of inflexibility, we address gaps in existing univariate studies by assessing the respective importance of sleep disturbances and chronotype distinctness in governing the emotional and cognitive impairment seen in individuals involved in shift working. We use Structural Equation Modelling (SEM) to capture the latent constructs governing sleep, mental health and cognition, and to model the complicated relationships between them. We hypothesise that individuals with greater chronotype distinctness will be more affected by shift work’s demands for circadian flexibility.

## METHODS AND MATERIALS

### participants

A cohort of 104 shift-working NHS nurses (*M_age_* = 36.29, *SD* = 9.03, female=85) were recruited from the EClocker study. Participants had no known history or current diagnosis of psychological or sleep disorder. The full inclusion criteria was (1) NMC registered NHS nurses working standard (day) shifts, or non-standard (rotating) shifts at participating NHS trusts, (2) aged ≥21 years. Exclusion criteria was (1) any mood disorder or sleep disorder diagnosis; alcohol / other substance dependence (excluding caffeine / nicotine) in the last 12 months, (2) illicit substance / psychostimulant use in the last 6 months, (3) pregnancy, (4) receiving a sleep intervention / treatment (including those which were research-led) or sleep-inducing medication (e.g., hypnotics, sedatives, melatonin) in the last 12 months. Two nurses dropped out during the data collection phase, making the final analysis sample n = 102.

### data acquisition / preparation

Participants in the EClocker study were invited to conduct a series of lifestyle questionnaires, as well as a battery of cognitive tasks. Here, we selected questionnaires targeting sleep, chronotype, and mental health symptoms, which included: the Pittsburgh Sleep Quality Index (PSQI) [20], Caen Chronotype Questionnaire (CCQ) [21], Mood Disorder Questionnaire (MDQ) [22], Patient Health Questionnaire (PHQ-8) [23], and Emotion Reactivity Scale (ERS) [24]. The cognitive battery was a standard battery designed by cognitive task platform Cognitron [25], involving four key tasks: Choice Reaction Time (CRT) to investigate attention and cognitive efficiency, Intra-Dimensional Extra-Dimensional task (IDED) for cognitive flexibility and attention switching, Learning Curves to investigate working memory, and Digit Span to assess attention, short-term and working memory. Cognitive tasks were assessed in a single time-point battery, with the exception of the CRT test which was assessed daily for 14 days. Cognitive test results were pre-processed in Python, producing a set of metrics for each task including total responses, median reaction time, error rate, and an overall summary statistic. Raw questionnaire data from the PSQI, CCQ, MDQ, PHQ-8, and ERS was scored and corroborated to ensure accuracy and to address missing data; blank entries were interpreted as non-symptomatic responses in all disorder-related questionnaires. Measurement variables were tested for skewness and kurtosis to ensure normality assumptions were met.

### measurement model

To accurately characterise the biological demands of shift work and the different aspects of associated symptom profiles, we wanted to capture the latent constructs assessed by each questionnaire without the limitation of selecting only one question from each. Hence, we proposed a measurement model to map multiple questionnaire elements onto their overarching construct, and used Confirmatory Factor Analysis (CFA) to confirm a factor structure for latent constructs underpinning sleep disturbances, chronotype distinctness, mood disorder, affective and somatic depressive symptoms, emotional reactivity, and cognitive impairment.

Observed variables chosen to represent each latent construct within the measurement model were based around previously validated constructs, such as recommended questionnaire sub-components. In some cases, we also used Exploratory Factor Analysis (EFA) for further refinement: from the cognitive metrics, the recommended components were the total number responses from the IDED task, error rate from the Digit Span task and median reaction time from the Learning Curves task. We also used EFA to separate the PSQI components specific to sleep quantification: quality, latency, duration and efficiency, from those more related to sleep health and medicine. We derived two factor-structures from the PHQ8 for depression, separating affective and somatic symptoms [26]. We used the three sub-components of the ERS questionnaire to capture emotional reactivity. The full measurement model prior to cleaning can be seen in the **Supplemental Figure**.

Factor loadings for observed variables were cleaned by removing weaker loadings (*r* < 0.55). The following variables with weaker associations to their construct were removed: sleep quality (*r* = 0.49) and latency (*r* = 0.36) from sleep; procrastination (*r* = 0.44), impairment (*r* = 0.47), and consistency of thinking (*r* = 0.49) from chronotype distinctness; talkativeness (*r* = 0.52) and hypersexuality (*r* = 0.54) from mood disorder; and altered speed (*r* = 0.55) from somatic depression. To optimise model complexity given the small sample size, factor loadings were further reduced to keep only the three strongest factor loadings for each construct. Covariance relationships were also included between working and focus, energy and activity, and insomnia and fatigue, to account for covarying errors due to similarities in their nature.

After the above refinement and cleaning of the model, the remaining components describing each latent construct were as follows: sleep disturbance: sleep duration (*r* = 0.82) and sleep efficiency (*r* = 0.75); chronotype distinctness: consistency of working (*r* = 0.58), mood (*r* = 0.73), and focus (*r* = 0.61); mood disorder: altered energy (*r* = 0.36), activity (*r* = 0.67), and sociability (*r* = 0.82). Two separate constructs were derived from the PHQ8 for depression, with affective depression: self-doubt (*r* = 0.66), low mood (*r* = 0.76) and apathy (*r* = 0.69); somatic depression: insomnia (*r* = 0.78), fatigue (*r* = 0.82), and altered appetite (*r* = 0.69). Emotional reactivity: emotional persistence (*r* = 0.79), arousal (*r* = 0.82) and sensitivity (*r* = 0.97). Cognition: IDED total responses (*r* = 0.82), Learning Curves median reaction time (*r* = 0.93), and Digit Span error rate (*r* = 0.87). The final cleaned measurement model can be seen in **Figure 1**.

**Figure 1.**
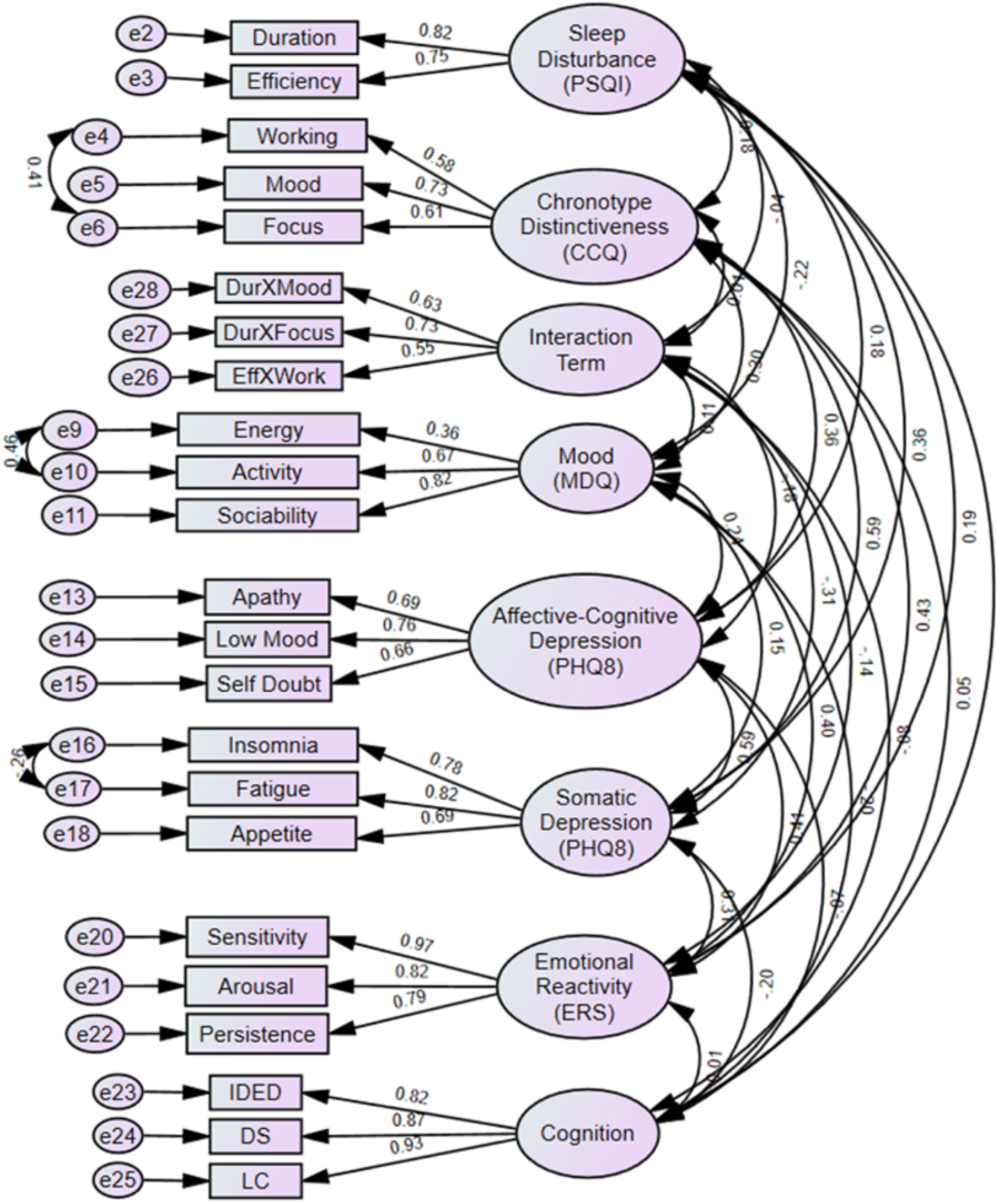
Cleaned measurement model.

Kaiser-Meyer-Olkin (KMO) and Bartlett’s tests were used to test for sampling adequacy and for the presence of sufficient underlying inter-variable correlations [27]; both were sufficient to justify complex modelling (KMO = 0.735; Bartlett’s: *χ*^2^(325) = 1199.46, *p* < 0.001).

The strength of the loadings between observed variable indicators and their latent constructs were tested for significance; constructs were tested for composite reliability and discriminant validity, and the overall fit of the cleaned measurement model was tested using *χ*^2^, *χ*^2^*/df*, CFI, TLI, and RMSEA. All observed variable indicators across the questionnaires and cognitive tests exhibited strong standardised loadings onto their constructs (0.54 *< r <* 0.96, *p* < 0.001), which all exhibited good composite reliability (> 0.72) and discriminant validity (*max*(*r*^2^) *< Var*). The overall measurement model demonstrated an excellent fit: *χ*^2^(199) = 227.001, *p* = 0.084, *χ*^2^*/df* = 1.14, CFI = 0.954, TLI = 0.967, RMSEA = 0.034, as indicated by the non-significant *χ*^2^, CFI and TLI values above 0.95, and RMSEA below 0.05.

### latent construct correlation analysis

Latent variables were computed according to factor loading-weighted sums of their contributing observed variables from the measurement model. We then computed Pearson correlations between each pair of variables in the day shift and rotating shift cohorts respectively, which revealed distinct relationships between the predictors (sleep disturbances and chronotype distinctness) and symptom profiles for each cohort (**Table 1**).

**Table 1:** Pearson correlation p-values between latent constructs including sleep disturbance (PSQI), chronotype distinctness (CCQ), mood disorder (MDQ), affective and somatic depression (PHQ8), emotional reactivity (ERS), and cognitive impairment **Note:** Significant correlations are indicated by *p < 0.05; **p < 0.001

|  | Sleep Disturbances (PSQI) | Chronotype Distinctness (CCQ) | Mood Disorder (MDQ) | Affective Depression (PHQ8) | Somatic Depression (PHQ8) | Emotional Reactivity (ERS) | Cognitive Impairment |
| --- | --- | --- | --- | --- | --- | --- | --- |
| Sleep Disturbances (PSQI) | 1.000 | 0.181 | 0.562 | 0.050* | 0.001** | 0.064 | 0.355 |
|  | 1.000 | 0.436 | 0.640 | 0.405 | 0.200 | 0.010** | 0.021* |
| Chronotype Distinctness (CCQ) |  | 1.000 | 0.235 | 0.125 | 0.002** | 0.115 | 0.908 |
|  |  | 1.000 | 0.048* | 0.015* | 0.003** | 0.012* | 0.754 |
| Mood Disorder (MDQ) |  |  | 1.000 | 0.195 | 0.262 | 0.259 | 0.254 |
|  |  |  | 1.000 | 0.243 | 0.803 | 0.383 | 0.510 |
| Affective Depression (PHQ8) |  |  |  | 1.000 | 0.004** | 0.035* | 0.294 |
|  |  |  |  | 1.000 | 0.001** | 0.117 | 0.729 |
| Somatic Depression (PHQ8) |  |  |  |  | 1.000 | 0.292 | 0.238 |
|  |  |  |  |  | 1.000 | 0.231 | 0.126 |
| Emotional Reactivity (ERS) |  |  |  |  |  | 1.000 | 0.744 |
|  |  |  |  |  |  | 1.000 | 0.025* |
| Cognitive Impairment |  |  |  |  |  |  | 1.000 |
|  |  |  |  |  |  |  | 1.000 |

In the day shift cohort, significant correlations were identified between sleep disturbance and affective (*p* = 0.050) and somatic depression (*p* = 0.001), and between chronotype distinctness and somatic depression (*p* = 0.003). Within the symptom profile, correlations were identified between affective and somatic depression (*p* = 0.004), and between affective depression and emotional reactivity (*p* = 0.035).

In the rotating shift cohort, significant correlations were identified between sleep disturbances and emotional reactivity (*p* = 0.010) and cognitive impairment (*p* = 0.021), and between chronotype distinctness and mood disorder (*p* = 0.048), affective depression (*p* = 0.015), somatic depression (*p* = 0.003), and emotional reactivity (*p* = 0.012). Within the symptom profile, correlations were identified between affective and somatic depression (*p* = 0.001), and between emotional reactivity and cognitive impairment (*p* = 0.025).

### structural equation modelling

We used Structural Equation Modelling (SEM) to model the above relationships between latent constructs for each cohort, modelling sleep disturbances and chronotype distinctness as predictors of symptom profiles. The two SEM models were constructed and evaluated using IBM SPSS AMOS [28].

For the day shift cohort (n = 45), we modelled sleep disturbances and chronotype distinctness as predictors of affective and somatic depressive symptoms and heightened emotional reactivity, according to significant relationships identified in the correlation analysis (**Figure 2**).

**Figure 2.**
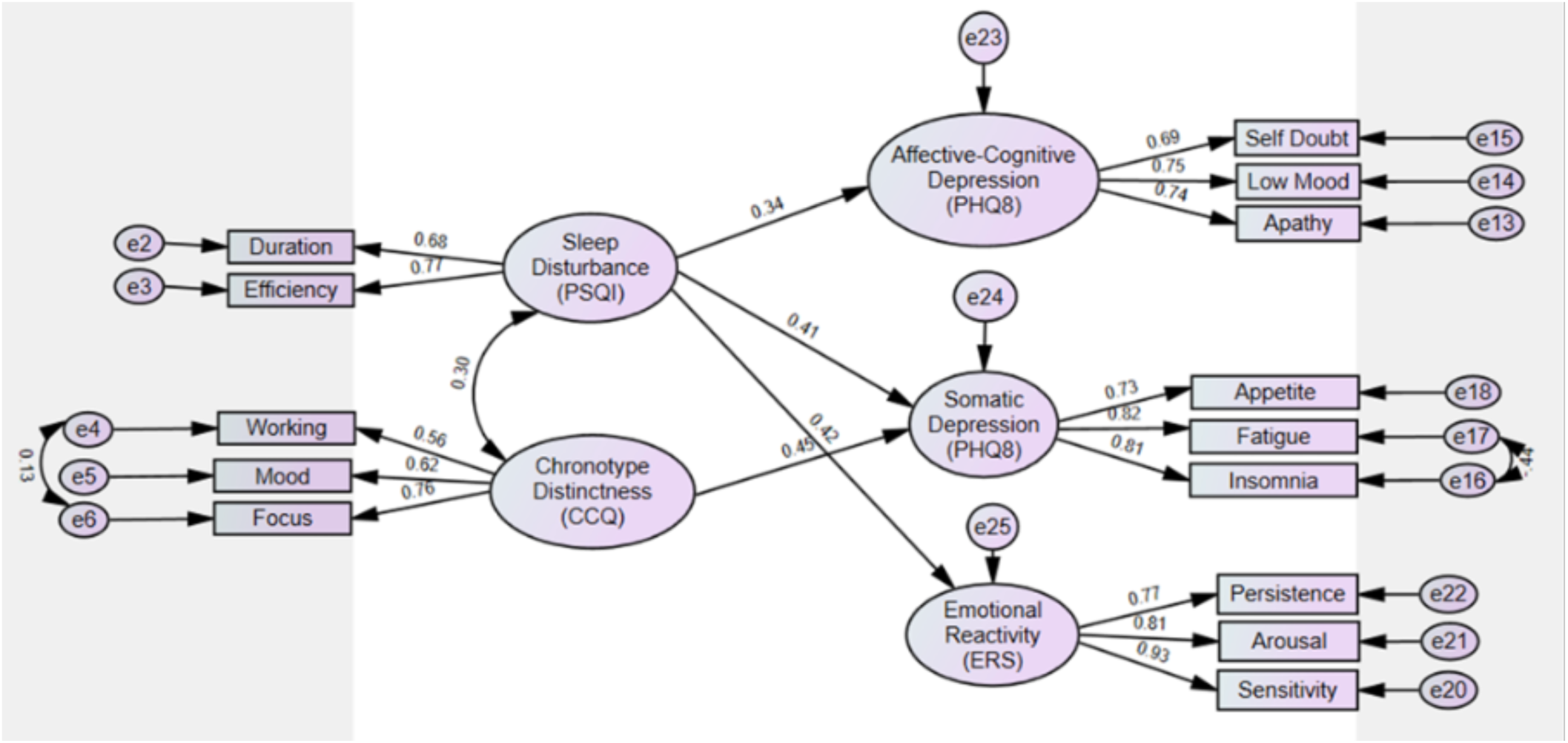
Day shift cohort structural equation model.

For the rotating shift cohort (n = 47), we modelled sleep disturbances as a predictor of cognitive impairment, and chronotype distinctness as a predictor of mood disorder, affective and somatic depressive symptoms, and heightened emotional reactivity (**Figure 3**).

**Figure 3.**
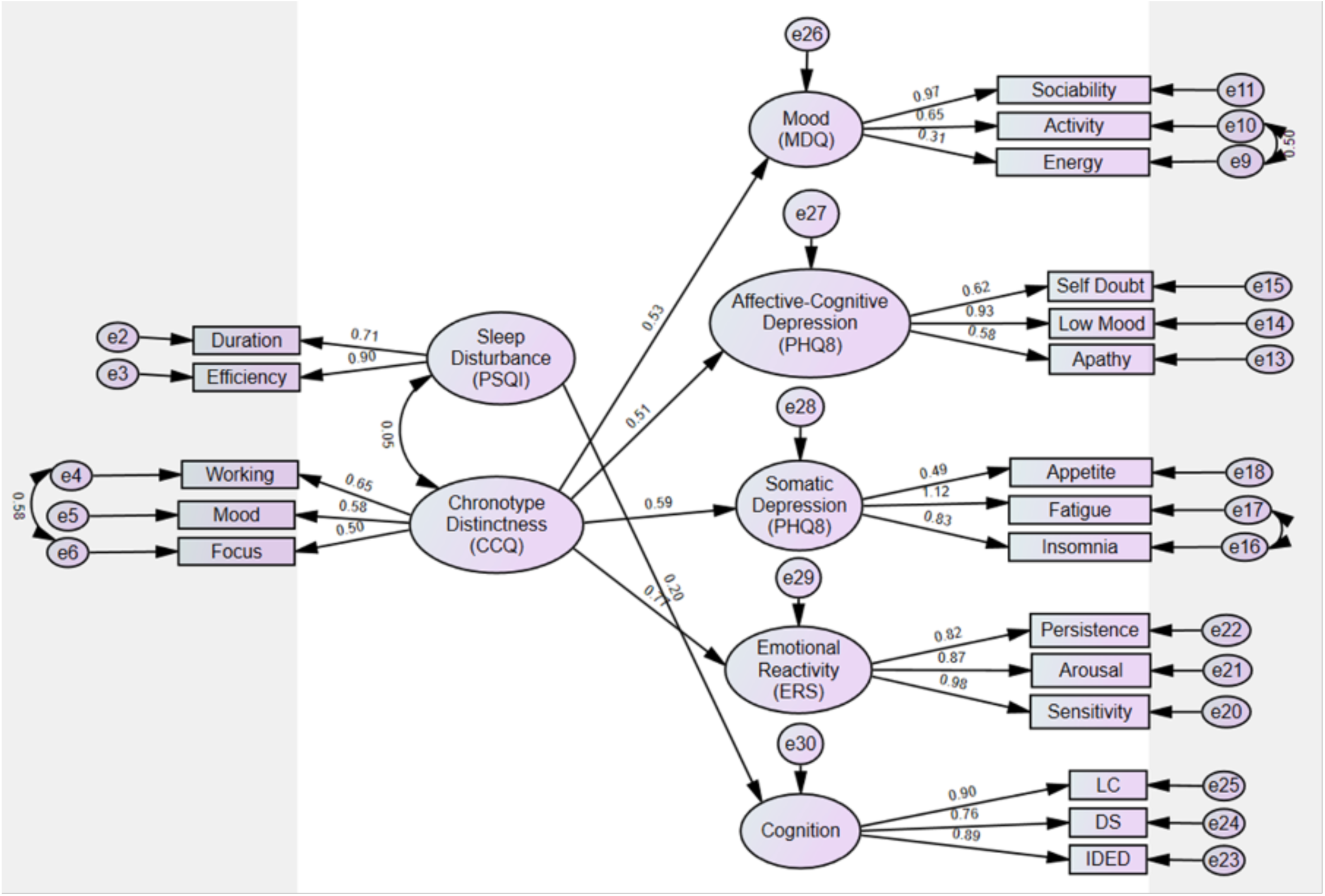
Rotating shift cohort structural equation model.

Both SEM models were tested for direct effects, significant paths, and overall model fit using *χ*^2^ test, *χ*^2^*/df*, CFI, TLI and RMSEA indices.

## RESULTS

### descriptive statistics

**Table 2** presents the descriptive statistics: mean scores, standard deviation, skewness and kurtosis for all observed variables used in the final measurement model. All variables fell within an acceptable range for skewness (< 2) and kurtosis (< 7), with the exception of the altered social behaviour item from the MDQ, which was partially skewed but considered to be within an acceptable range given the healthy nature of the cohort. All variables were sufficiently normally distributed to be included in subsequent analysis.

**Table 2:**
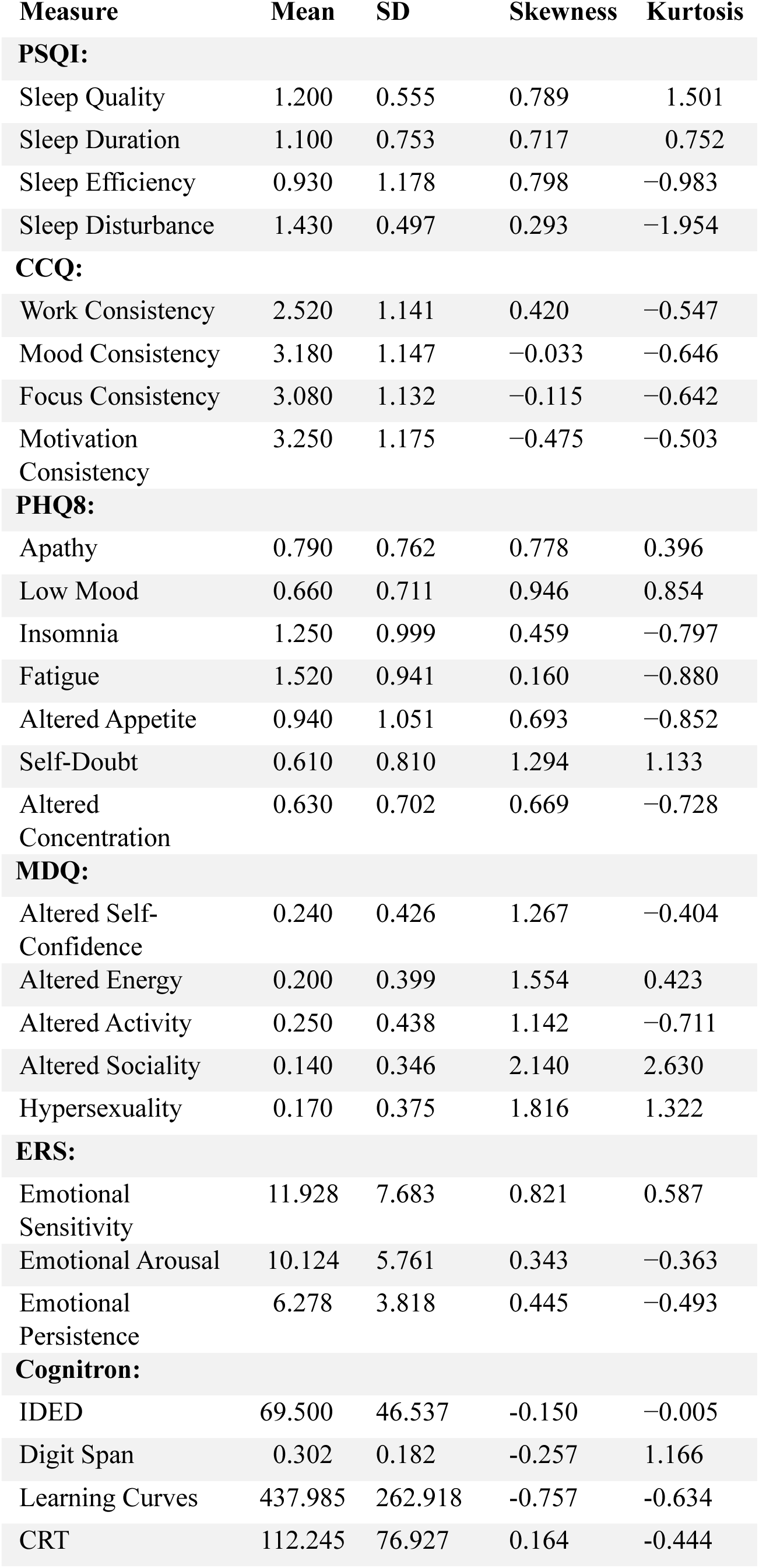
Descriptive statistics for key questionnaire components and cognitive task measures.

| Measure | Mean | SD | Skewness | Kurtosis |
| --- | --- | --- | --- | --- |
| **PSQI:** |  |  |  |  |
| Sleep Quality | 1.200 | 0.555 | 0.789 | 1.501 |
| Sleep Duration | 1.100 | 0.753 | 0.717 | 0.752 |
| Sleep Efficiency | 0.930 | 1.178 | 0.798 | −0.983 |
| Sleep Disturbance | 1.430 | 0.497 | 0.293 | −1.954 |
| **CCQ:** |  |  |  |  |
| Work Consistency | 2.520 | 1.141 | 0.420 | −0.547 |
| Mood Consistency | 3.180 | 1.147 | −0.033 | −0.646 |
| Focus Consistency | 3.080 | 1.132 | −0.115 | −0.642 |
| Motivation Consistency | 3.250 | 1.175 | −0.475 | −0.503 |
| **PHQ8:** |  |  |  |  |
| Apathy | 0.790 | 0.762 | 0.778 | 0.396 |
| Low Mood | 0.660 | 0.711 | 0.946 | 0.854 |
| Insomnia | 1.250 | 0.999 | 0.459 | −0.797 |
| Fatigue | 1.520 | 0.941 | 0.160 | −0.880 |
| Altered Appetite | 0.940 | 1.051 | 0.693 | −0.852 |
| Self-Doubt | 0.610 | 0.810 | 1.294 | 1.133 |
| Altered Concentration | 0.630 | 0.702 | 0.669 | −0.728 |
| **MDQ:** |  |  |  |  |
| Altered Self-Confidence | 0.240 | 0.426 | 1.267 | −0.404 |
| Altered Energy | 0.200 | 0.399 | 1.554 | 0.423 |
| Altered Activity | 0.250 | 0.438 | 1.142 | −0.711 |
| Altered Sociality | 0.140 | 0.346 | 2.140 | 2.630 |
| Hypersexuality | 0.170 | 0.375 | 1.816 | 1.322 |
| **ERS:** |  |  |  |  |
| Emotional Sensitivity | 11.928 | 7.683 | 0.821 | 0.587 |
| Emotional Arousal | 10.124 | 5.761 | 0.343 | −0.363 |
| Emotional Persistence | 6.278 | 3.818 | 0.445 | −0.493 |
| **Cognitron:** |  |  |  |  |
| IDED | 69.500 | 46.537 | −0.150 | −0.005 |
| Digit Span | 0.302 | 0.182 | −0.257 | 1.166 |
| Learning Curves | 437.985 | 262.918 | −0.757 | −0.634 |
| CRT | 112.245 | 76.927 | 0.164 | −0.444 |

Despite the target demographic being healthy individuals, the questionnaires detected high rates of mental health disorder within the cohort. The prevalence of each disorder according to typical diagnostic thresholds were as follows: sleep disorder: 77.6% (PSQI global score ≥5), moderate to severe depression: 36.3% (PHQ8 global score ≥8).

### day shift model results

The day shift model demonstrated a good fit (n=49): *χ*^2^(70) = 90.114, p = 0.053, *χ*^2^*/df* = 1.29, CFI = 0.912, TLI = 0.885, RMSEA = 0.077. Given the small sample size, these fit statistics indicate a strong model, particularly the non-significant *χ*^2^ result. Significant paths were found between sleep disturbances and somatic depression (*p* = 0.03), sleep disturbances and emotional reactivity (*p* = 0.03), and between chronotype distinctness and somatic depression (*p* = 0.025). Hence, sleep disturbance emerged as the key driver of adverse symptoms in day shift workers.

### rotating shift model results

The rotating shift work model demonstrated a moderate fit: *χ*^2^(161) = 222.77, *p* = 0.001, *χ*^2^*/df* = 1.38, CFI = 0.868, TLI = 0.827, RMSEA = 0.086. Sample size and model complexity are known to impact model fit statistics, particularly TLI and RMSEA [29]. Despite this effect, the strength of paths within the model provide a strong basis for this model. Significant paths were found between chronotype distinctness and mood disorder (*p* = 0.004), affective depression (*p* = 0.006), somatic depression (*p* = 0.004), and emotional reactivity (*p* < 0.001). Hence, individual chronotype distinctness emerged as the key driver of adverse symptoms in rotating shift workers, increasing the risk of mood and affective disorders, and exacerbating the effects of sleep disturbance on cognitive function.

## DISCUSSION

This study found significant links between the physiological demands of shift working and detrimental mental health and cognitive outcomes in healthcare workers. A high prevalence of mental disorders was observed within the cohort of NHS nurses, with all experiencing at least one clinically relevant disorder, including sleep disorders in 77.6% and moderate to severe depressive symptoms in 36.3%. Significant cognitive impacts were also observed, particularly amongst those participating in rotating shifts, indicating that the disruption may extend beyond personal affective experiences into disrupted occupational efficiency and even safety. Isolating two drivers of biological insult associated with healthcare and shift work: sleep disturbance and chronotype distinctness, revealed that both play important but distinct roles in governing symptom profiles, depending on the nature of the work schedule.

The high rates of disturbances within the cohort highlight the significant psychiatric risk posed by the demands of healthcare-based work, and the need for remediative interventions, reflecting similar findings across the healthcare sector [11]. Further to optimising working conditions to reduce the biological insult of shift-based work (e.g., through shift design), interventions should consider addressing the psychological demands on healthcare workers (e.g., frequent exposure to stressful events in nurses) [30].

For individuals involved in day shift work, sleep disturbances consistently emerged as the leading cause of health issues, which were centred around somatic depressive symptoms and increased emotional reactivity. This increase in physical symptoms of depression is likely to be related to increased fatigue and sleep pressure [31]. Separating the affective and somatic symptoms of depression revealed that the high rates of depression-like symptoms within the cohort are more related to physical fatigue than psychological depressive symptoms such as anhedonia; fatigue is often linked with adverse symptoms in healthcare workers [32]. Interventions for day shift workers should aim to target sleep deprivation, by educating employees about improving sleep hygiene or by increasing sleep opportunity through shift re-design. Common practices such as shift-swapping and overtime, whilst allowing organisations to ensure delivery, may contribute to the impairments identified by this study, and should hence be entered into with better awareness of the associated risks. Improved understanding may also contribute to an increased sense of agency over individuals’ mental health, something that has been shown to improve depressive symptoms [33]. Research into attitudes surrounding depression has found that, whilst the belief that depressive symptoms are caused by an underlying chemical imbalance may reduce personal blame, it also reduces personal accountability and empowerment compared to an understanding of potential biopsychosocial causes that could be targeted by intervention [34].

For healthcare workers involved in rotating shift work, though the nature of their occupation is the same, they are exposed to a completely different environment due to schedule variability, disrupted or restricted light exposure, and inconsistently timed biological demands. For this cohort, the impact of short-term sleep disturbances appeared to be eclipsed by the effects of the physiological insult caused by this altered environment, with individual chronotype distinctness governing the extent of detrimental symptoms, further validating chronotype distinctness as a marker of inflexibility to altered routines. Symptom profiles within the rotating shift cohort included affective and somatic depressive symptoms and emotional reactivity, and extended to include mood disorder and even cognitive impairment. Although sleep health has been considered, the literature highlights the cascading impacts of circadian health, where sleep is only one component. Future research should adopt a more holistic approach, and consider how the remaining circadian processes (cardiovascular, digestive, immunological etc.) are differentially affected in fast rotating shift workers. Interventions for such workers should target both sleep disturbances and circadian dysregulation, through interventions such as targeted light entrainment.

Whilst chronotype distinctness emerged as a key indicator of individual sensitivity to rotating shift work, the nature of this natural flexibility is not yet thoroughly understood. Here, we have considered distinctness as a fixed, trait-based indicator of flexibility, in which case more flexible individuals could be considered more robust to the demands of rotating shift work than others. However, it is possible that an individual’s distinctness may vary across their lifespan, or in response to other environmental factors, in which case targeted interventions could aim to increase flexibility. Irrespective of this, inflexibility to the demands of shift working should be considered as a key risk factor for developing adverse psychiatric symptoms, meriting close monitoring and mediative measures for at-risk individuals.

These initial findings highlight the importance of considering the full symptom profile when assessing the effects of shift working; future studies should aim to recruit a larger cohort, perhaps across a range of employment sectors to further investigate the role of distinctness in different shift work settings. Longitudinal studies could also be used to investigate the exacerbation of symptoms with chronic exposure to the shift work system.

## CONCLUSION

We found that adverse symptom profiles in healthcare workers are led by sleep disturbances for those involved in day shifts, but governed increasingly by individual chronotype distinctness when variable schedules are introduced. We also found that these symptom profiles, restricted to affective disorders and mild emotional dysregulation in day shift workers, extend to include mood disorders and cognitive impairment in rotating shift workers. Hence, we identify chronotype distinctness as a key risk factor for detrimental effects caused by variable shift working. Future work should seek to determine which types of variable schedule cause the most significant circadian insult, and to investigate the mediative role of chronotype distinctness on the effects of schedule variability.

## CONFLICTS OF INTEREST

AH is cofounder and codirector of H2 Cognitive Designs, a company that licenses online assessment technology for research and healthcare purposes. AH is also founder and director of Future Cognition Ltd, a company that develops cognitive assessment technology for third parties. PJH is also cofounder and codirector of H2 Cognitive Designs. The remaining authors declare that the research was conducted in the absence of any commercial or financial relationships that could be construed as a potential conflict of interest. All authors have completed the Unified Competing Interest form (available on request from the corresponding author) and declare: no support from any organisation for the submitted work; no financial relationships with any organisations that might have an interest in the submitted work in the previous years, no other relationships or activities that could appear to have influenced the submitted work.

## FUNDING STATEMENT

This paper represents independent research [part] funded by the National Institute for Health and Care Research (NIHR) Maudsley Biomedical Research Centre at South London and Maudsley NHS Foundation Trust and King’s College London. The views expressed are those of the author(s) and not necessarily those of the NIHR or the Department of Health and Social Care.

## DECLARATION

The Corresponding Author has the right to grant on behalf of all authors and does grant on behalf of all authors, an exclusive licence (or non-exclusive for government employees) on a worldwide basis to the BMJ Publishing Group Ltd to permit this article (if accepted) to be published in BMJ editions and any other BMJPGL products and sublicenses such use and exploit all subsidiary rights, as set out in our licence. The authors affirm that the manuscript is an honest, accurate, and transparent account of the study being reported; that no important aspects of the study have been omitted; and that any discrepancies from the study as planned have been explained.

## ETHICAL APPROVAL

This study involves human participants and was approved by the Research Ethics Committee at King’s College London (HR-19/20-17792). The study was also approved by the Health Research Authority (HRA) and all participating NHS Trusts, Integrated Research Application System (IRAS) (289592). NHS organisations operated as Participant Identification Centres (PICs). Participants gave informed consent to participate in the study before taking part.

## Data Availability

All data produced in the present work are contained in the manuscript

---

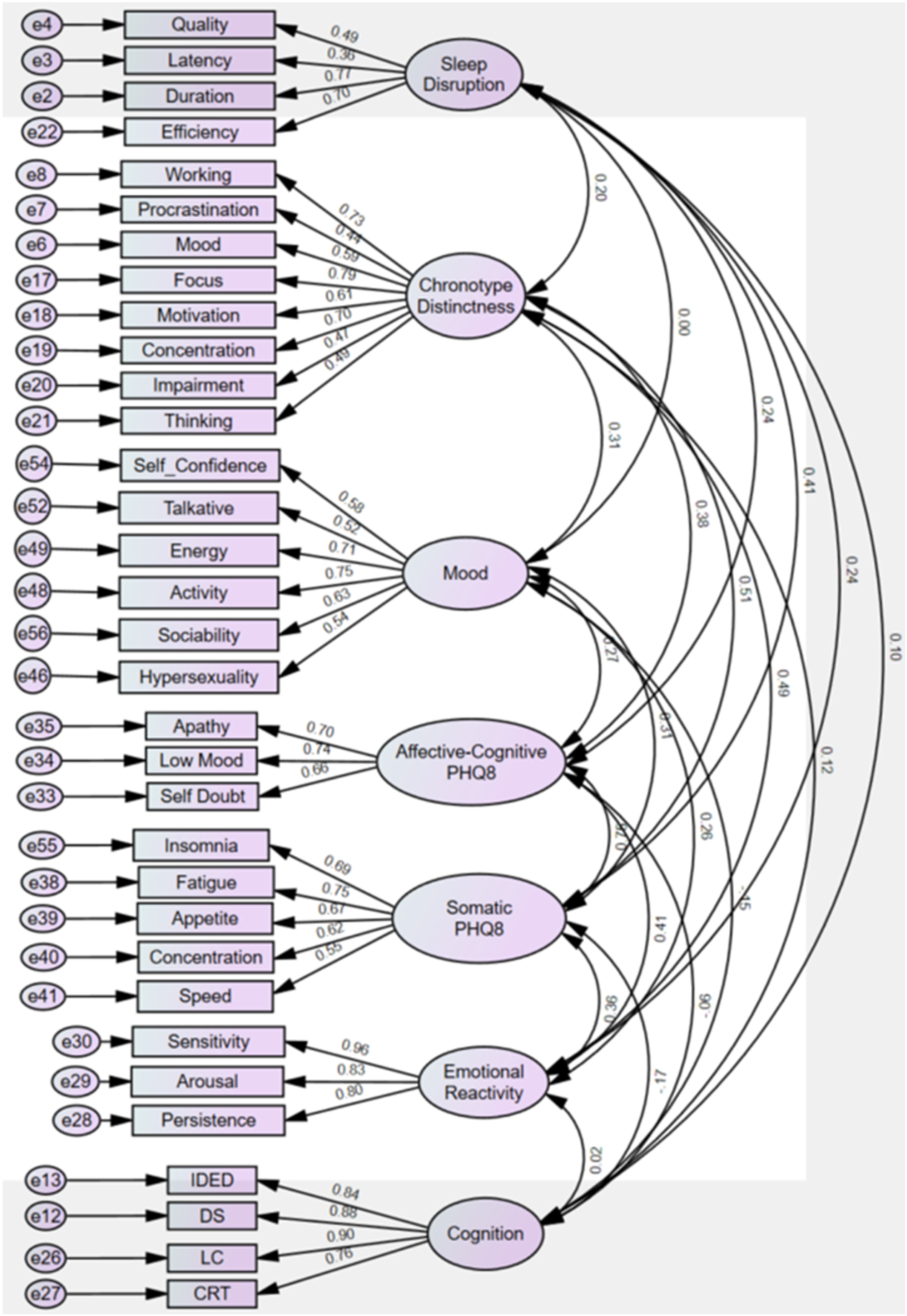
Supplemental Figure. Full measurement model.

## Notes

### Author Declarations

The Research Ethics Committee of King's College London gave ethical approval for this work.

## REFERENCES

[1] Barkoukis T. Therapy in sleep medicine. Elsevier 2012.

[2] Office for National Statistics. The healthcare workforce across the uk: 2024 [online]. 2024. https://www.ons.gov.uk/peoplepopulationandcommunity/he althandsocialcare/healthcaresystem/articles/thehealthcarewo rkforceacrosstheuk/2024 (accessed 7 October 2025).

[3] Reynolds AC, Ferguson SA, Appleton SL, et al. Prevalence of probable shift work disorder in non-standard work schedules and associations with sleep, health and safety outcomes: A cross-sectional analysis. Nat. Sci. Sleep 2021; 13:683–693.

[4] Hastings MH, Maywood ES, Brancaccio M. Generation of circadian rhythms in the suprachiasmatic nucleus. Nat. Rev. Neurosci. 2018; 19(8): 453–469.

[5] Michel S, Johanna HM. From clock to functional pacemaker. Eur. J. Neurosci. 2020; 51(1):482–493.

[6] Duffy JF, Wright KP. Entrainment of the human circadian system by light. J. Biol. Rhythms 2005; 20(4):326–338.

[7] McGovern A, MIT Lincoln Laboratory. New technologies tackle brain health assessment for the military [online]. 2025. https://news.mit.edu/2025/new-technologies-tackle-brain-health-assessment-for-military-0825 (accessed 11 October 2025).

[8] Barger, LK, Ayas NT, Cade BE, et al. Impact of extended-duration shifts on medical errors, adverse events, and attentional failures. PLoS Med. 2006; 3(12):487.

[9] Hickman R, D’Oliveira T, Davies A, et al. Monitoring daily sleep, mood, and affect using digital technologies and wearables: A systematic review. Sensors (Basel) 2024; 24(14):4701.

[10] Akerstedt T. Shift work and disturbed sleep/wakefulness. Occup. Med. (Lond.) 2003; 53(2): 89–94.

[11] D’Oliveira TC, Anagnostopoulos A. The association between shift work and affective disorders: A systematic review. Chronobiol. Int. 2021; 38(2):182–200.

[12] Kaliyaperumal D, Elango Y, Alagesan M, et al. Effects of sleep deprivation on the cognitive performance of nurses working in shift. J. Clin. Diagn. Res. 2017; 11(8):CC01– CC03.

[13] Dai C, Qiu H, Huang Q, et al. The effect of night shift on sleep quality and depressive symptoms among chinese nurses. Neuropsychiatr. Dis. Treat. 2019; 15: 435–440.

[14] Kalmbach DA, Pillai V, Roth T, et al. The interplay between daily affect and sleep: a 2-week study of young women. J. Sleep Res. 2014; 23(6):636–645.

[15] Li Y, Androulakis IP. Light entrainment of the SCN circadian clock and implications for personalized alterations of corticosterone rhythms in shift work and jet lag. Sci. Rep. 2021; 11(1):17929.

[16] Cheng W-J, Puttonen S, Vanttola P, et al. Association of shift work with mood disorders and sleep problems according to chronotype: a 17-year cohort study. Chronobiol. Int. 2021; 38(4):518–525.

[17] Dosseville F, Laborde S, Lericollais R. Validation of a chronotype questionnaire including an amplitude dimension. Chronobiol. Int. 2013; 30(5): 639–648.

[18] Marcoen N, Vandekerckhove M, Neu D, et al. Individual differences in subjective circadian flexibility. Chronobiol. Int. 2015; 32(9):1246–1253.

[19] Hickman R, Lai Jie D, Shergill S, et al. Validation of the caen chronotype questionnaire: Exploring the added value of amplitude and correlations with actigraphy. Chronobiol. Int. 2025; 42(3):378–391.

[20] Buysse DJ, Reynolds CF, Monk TH, et al. The pittsburgh sleep quality index: a new instrument for psychiatric practice and research. Psychiatry Res., 1989; 28(2):193–213.

[21] Dosseville F, Laborde S, Lericollais R. Validation of a chronotype questionnaire including an amplitude dimension. Chronobiol. Int. 2013; 30(5): 639–648.

[22] Hirschfeld RM, Williams JB, Spitzer RL, et al. Development and validation of a screening instrument for bipolar spectrum disorder: the mood disorder questionnaire. Am. J. Psychiatry. 2000; 157(11):1873– 1875.

[23] Kroenke K, Strine TW, Spitzer RL, et al. The PHQ-8 as a measure of current depression in the general population. J. Affect. Disord. 2009; 114(1-3):163–173.

[24] Nock MK, Wedig MM, Holmberg EB, et al. The emotion reactivity scale: development, evaluation, and relation to self-injurious thoughts and behaviors. Behav. Ther. 2008; 39(2): 107–116.

[25] Cognitron. Cognitron online cognitive assessment technology [online]. 2025. https://www.cognitron.co.uk/ (accessed 11 October 2025)

[26] Vu LG, Le LK, Dam AVT, et al. Factor structures of patient health questionnaire 9 instruments in exploring depressive symptoms of suburban population. Front. Psychiatry. 2022; 13:838747.

[27] IBM. Kmo and bartlett’s test [online]. 2025. https://www.ibm.com/docs/en/spss-statistics/28.0.0?topic=detection-kmo-bartletts-test (accessed 12 October 2025).

[28] IBM Corp. Ibm spss amos (version 29.0) [online]. 2024. https://www.ibm.com/products/structural-equation-modeling-sem (accessed 12 October 2025).

[29] Shi D, Lee T, Maydeu-Olivares A. Understanding the model size effect on SEM fit indices. Educ. Psychol. Meas. 2019; 79(2):310–334.

[30] Weinberg A, Creed F. Stress and psychiatric disorder in healthcare professionals and hospital staff. Lancet. 2000; 355(9203):533–537.

[31] Reichert CF, Maire M, Schmidt C, et al. Sleep-wake regulation and its impact on working memory performance: The role of adenosine. Biology (Basel). 2016; 5(1):11.

[32] Owens JA. Sleep loss and fatigue in healthcare professionals. J. Perinat. Neonatal Nurs. 2007; 21(2):92–100.

[33] Lin J, Yang X, Li H, et al. Enhancing agency in individuals with depressive symptoms: The roles of effort, outcome valence, and its underlying cognitive mechanisms and neural basis. Depress. Anxiety. 2024; 2024(1):3135532.

[34] Deacon BJ, Baird GL. The chemical imbalance explanation of depression: Reducing blame at what cost? J. Soc. Clin. Psychol. 2009; 28(4):415–435

